# COVIDPEN: A Novel COVID-19 Detection Model using Chest X-Rays and CT Scans

**DOI:** 10.1101/2020.07.08.20149161

**Authors:** Amit Kumar Jaiswal, Prayag Tiwari, Vipin Kumar Rathi, Jia Qian, Hari Mohan Pandey, Victor Hugo C. Albuquerque

## Abstract

The trending global pandemic of COVID-19 is the fastest ever impact which caused people worldwide by severe acute respiratory syndrome (SARS)-driven coronavirus. However, several countries suffer from the shortage of test kits and high false negative rate in PCR test. Enhancing the chest X-ray or CT detection rate becomes critical. The patient triage is of utmost importance and the use of machine learning can drive the diagnosis of chest X-ray or CT image by identifying COVID-19 cases. To tackle this problem, we propose COVIDPEN - a transfer learning approach on Pruned EfficientNet-based model for the detection of COVID-19 cases. The proposed model is further interpolated by post-hoc analysis for the explainability of the predictions. The effectiveness of our proposed model is demonstrated on two systematic datasets of chest radiographs and computed tomography scans. Experimental results with several baseline comparisons show that our method is on par and confers clinically explicable instances, which are meant for healthcare providers.

## I. Introduction

World Health Organization (WHO) declared COVID-19 as global pandemic in March, 2020. It has caused catastrophic damage globally with more than four million confirmed cases and, more than 280 thousand deaths reported at the time of this writing. Countries across the world have taken stringent measures to flatten the curve corresponding to new cases and to slow down the rate of spread of the virus. Such efforts would buy the scientists more time for vaccine development. It is also imperative to improve diagnostic techniques to facilitate early detection of the virus. This is also required in order to reduce the time to declare the test results as well as to enhance the accuracy of the test. With the massive testing everyday, achieving the aforementioned goals is need of the hour.

As of now, there are three main methods to detect COVID-19:

- Polymerase Chain Reaction (PCR)
- Chest Computed Tomography (CT)
- Detection of Antibodies

PCR is based on the respiratory samples obtained by sputum, nasopharyngeal swab, etc. There are numerous issues in utilizing this technique at present. It takes a lot of time to produce the massive test kits and to output the available result. Also, the positivity rate is only 63% [1]. In [2], the authors point out that the false negative result produced by PCR is due to the inappropriate way of extraction of nucleic acid from clinical substrates and lesser cellular material for identification. This increases the risk of inaccurate and delayed diagnosis leading to more people being effected from the individual under test.

CT has been considered as a promising method to assist the final diagnosis, in particular, when the amount of test kits cannot meet the testing requirement. A study in Wuhan suggests that the CT method has been significantly more sensitive than PCR at early stages. These images may help in early detection of the disease. The problem, however, in this approach is that the images from COVID-19 infections are similar to those of different types of viral pneumonia that relate to inflammatory lung diseases. Hence, the CT images demonstrate similar patterns like bilateral, multifocal, opacities, mainly in the lower lobes, in the early stage and pulmonary consolidation in the late stages [3]. This increases the chances of a false negative or false positive.

Significant researches suggest notable changes in the CT scans of infected individuals with respect to time. In the study performed on 121 symptomatic patients in four centres in China, it was reported that in the early stages, the CT scans were relatively normal. After the onset of symptoms, the findings become more prominent with greater total lung involvement, crazy-paving patterns and reverse halo signs. For instance, bilateral lung involvement was visualized in only 28% of early patients, increased to 76% in the medieval period and rose to 88% in later stages. Hence, CT scans can be extremely useful to detect the infection in at least intermediate stages if not initial ones. In [4], the authors acknowledge the presence of ground glass and consolidative opacities in CT scans of infected patients, confirming four other researches. They also pointed towards inability of CXR images to help visualization of such manifestations. The authors also mention infected cases with no chest CT abnormalities that raises the challenge for detection and calls for improved techniques.

In the present work, an autonomous COVID-19 detection technique has been proposed utilizing Artificial Intelligence to analyze CT scans. Deep learning techniques have been deployed to study the images and classify them as COVID-19 positive or negative. Several papers have highlighted the usage of deep learning for detecting lung diseases like pneumonia. In [5], the authors devised a 3D-deep learning framework for identification of COVID-19. It extracts both 2D local and 3D global representative features. The results show high specificity of 96% in detecting COVID-19. The model can delineate pneumonia and other similar diseases from COVID-19 to quite a good extent saying a lot about the potential of the framework.

We summarise our contributions as follows:

- We propose COVIDPEN - a transfer learning approach adapted on **P**runed **E**fficient**N**et [6] model for automatic detection of COVID-19 disease.
- The feasibility of our proposed method on a real COVID-19 chest X-ray / CT scans dataset demonstrates its effectiveness.

The paper has been structured as follows. The next section discusses some related work in the area of medical image analysis using machine learning tools. Section III presents the proposed COVID-19 detection model. Section IV outlines the experimental evaluation obtained and performance analysis of this approach. The last section concludes the paper.

## II. Related Work

Analysing chest radiographs and CT scans has been seen as a manual task for medical practitioners / experts due to significant time effort. However, medical imaging has been advanced to current trend of technologies such as predictive modeling using machine learning and vision-driven tools (computer vision). Past work demonstrated few methods [7], [8] to diagnose diseases by means of providing decisions [9] and exploratory insights to X-ray images. This formulates as a precise use case for the Deep Learning framework.

Deep Learning (DL) algorithms are widely applicable on high-dimensional datasets, for instance, images. In particular, with the increasing computation power (GPU), grasp of error propagation and, improved optimization methods, more and more practical problems can be addressed by DL. It allows multiple layers to stack together enabling it to learn complex representations. These representations placed on layers correspond to different abstract levels and eventually collaborate to implement the objective task (e.g., classification) at the last layer. The process equates to the linear combination of all the representations (features).

Many researchers investigated the application of DL on several medical imaging tasks. Earlier work such as [10], [11] introduced a neural network approach to identify the lymph node under low contrast background composition in a diagnostic tasks and the classification of lung disease using deep neural network including the identification of thoraco-abdomincal lymph. They also made an extensive comparative analysis with baseline neural network models which showed the effectiveness of their method where recall of 85% with the rate of 3 false positives patient. A semi-supervised learning approach was adapted on a deep convolutional neural network to identify the metastases cancer cells in histopathological scans [12]. Their proposed model is generalizable on smaller set of image sample and shown to be effective (in terms of performance). A recent work [13] on magnetic resonance imaging (MRI) used CNN-based model to analyze the samples from spinal lumber images. Also, their CNN model shown to be effective in creation of Pfirrmann grading of spinal lumber MRIs. In [14], an initial model that pre-trained on ImageNet was developed and then fed into a classifier for further classification.

A deep neural network was designed for the chest X-ray14 dataset, claiming to have the capability to identity 14 categories of diseases with high efficiency [15]. In [16], Mask-RCNN was deployed for pneumonia identification. The proposed method follows residual proposal network alongwith pixel-wise segmentation of augmented pulmonary images. A follow-up work on the similar line of research [17] for pediatric pneumonia detection used a residual network composed of 49 convolutional layers which has one 2D-global average pooling layer and two dense layers. In [18], two CNN based models were used: AlexNet-based convolutional neural network and ResNet18. The first was used for the classification of lung patches and the the deep neural network-based model on ResNet18 were used to generate the hidden region of the lung image. Th outputs are the initial segmentation and reconstruction via ensembling models.

Some researchers have proposed techniques based on DL to diagnose for COVID-19 based on CT images. We discuss some of these works below.

A ResNet-50 based CNN model was proposed by Fu et. al. [19] to classify CT scans into five categories: COVID-19, non-COVID-19, normal, pulmonary tuberculosis, and bacterial pneumonia. The pre-trained residual network (ResNet) model was initially trained on ImageNet where tuning were performed on the weights of the last three convolutional layers alongwith the last fully connected layer. The accuracy on test data were reported to be 99.4%, 98.8%, 98.5%, 98.3%, and 98.6% for normal lungs, COVID-19, non-COVID-19 viral pneumonia, bacterial pneumonia, and pulmonary tuberculosis respectively. In [20], the authors used five state of the art CNN models: VGG19, Mobile Net, Inception, Xception, and Inception ResNet v2 for the classification of X-Ray images into one of the diagnosis label such as pneumonia, COVID-19, and normal. Transfer learning was used to be able to develop a good model trained on a small image dataset as the authors worked on a dataset with 224 COVID-19 positive images. Best results were obtained with VGG-19 and Mobile Net with accuracy of 98.75% and 97.40 % respectively while detecting COVID-19 only. Highest overall accuracy was reported as 93.48%. Also, it was shown that Mobile Net outperformed VGG19 on the basis of specificity and proved to be a relatively better model. Wang et.al. [1] suggested a deep neural network-driven model for prediction of Covid-19 which is termed as Covid-Net trained on the dataset COVIDx. The architecture built on a PEPX design pattern was first pre-trained on ImageNet and also utilized data augmentation. An accuracy of 92.4% was reported with 80% sensitivity for COVID-19 and very few false positive COVID-19 detections. This work [21] suggests the use of test-time augmentation (TTA) to improve the model’s accuracy. TTA produces transformed versions of images for prediction and the paper shows that even simple TTA techniques like rotation can improve the results significantly. The authors used TTA with two deep learning models (UNet and Mask R-CNN) built to segment nuclei in microscopic images. The results show higher segmentation accuracy.

## III. The Proposed Model

### A. Problem Formulation

An overview of a classification task we tackle in this work is outlined. In this paper, we consider a task of identifying COVID-19 disease which is a binary classification, where the input to COVIDPEN is a chest X-ray or CTs image *I*_*x*_ and the model outputs a binary label *P*_*y*_ ∈ {*positive, negative*} delineating whether the coronavirus prediction is positive or negative. For training optimization on imbalanced dataset, we use the Focal loss [22]

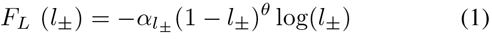

where *l*_*±*_ and *α*_*±*_ are defined as

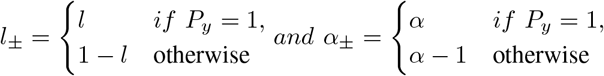

where *P*_*y*_ ∈ {− 1, 1} represents a binary label which contains positive and negative classes of COVID-19 bacterial disease labels, *l* ∈ [0, 1] represents the detected probability class label of model *l* = 1, and the parameter *α* ∈ [0, 1] depicts threshold which is to adjust the needfulness of positive and negative labeled disease samples, whereas the non-negative tuning parameter *θ* smoothly balances the rate at which sample instances are down weighted. However, when *λ* equals 0, the focal loss become sigmoid cross-entropy loss.

The usefulness of such loss function is that it corrects samples with implicit values which fits hard classification, which means the loss function is down-weighted for detected samples so that their role to the overall loss is comparatively small.

### B. Modelling

This section details the proposed model shown in Fig. 1 which is based on the EfficientNet model [6]. We employ several convolutional neural network (CNN) based models such as VGG19 [23], ResNet [24] variants and DenseNet [25] which in particular is developed for image classification and other computer vision tasks. However, these CNNs differs in a way as it captures the temporal and spatial dependencies (or features) from the input image and optimizes the number of parameters. We found residual networks [24] to perform better for our task, which is identifying a frontal-view X-ray image that outputs a binary prediction label to whether it is positive or negative. However, from performance perspective of the classifier ResNet50, it stands as a significantly best model among others but it lacks an inherent way to encode image features due to unbalanced COVID-19 chest X-ray samples. We examine the performance of other CNN-based models across different evaluation measures and found EfficientNet [6] to be a better alternative that imposes a symmetry between all convolutional layers. However, EfficientNet-B0 does not perform well as ResNet does, and we fine-tuned [6] which gives the classifier a significant improvement. Although, the test accuracy of fine-tuned EfficientNet on improvement is lower than ResNet50 which posed a major challenge for such deep neural networks to reason themseleves. To overcome the performance issue on less samples of image data, we adopted transfer learning mechanism to avoid overfitting, error and time for pseudo-labelling. The transfer learning technique is an effective approach to inherently ship the extracted knowledge from a network (ImageNet) trained on large collection of images with diverse features (such as color, shape etc.). The major importance of transfer learning approach is to enrich the existing parameters such as convolution weights which is trained on ImageNet.

**Fig. 1:**
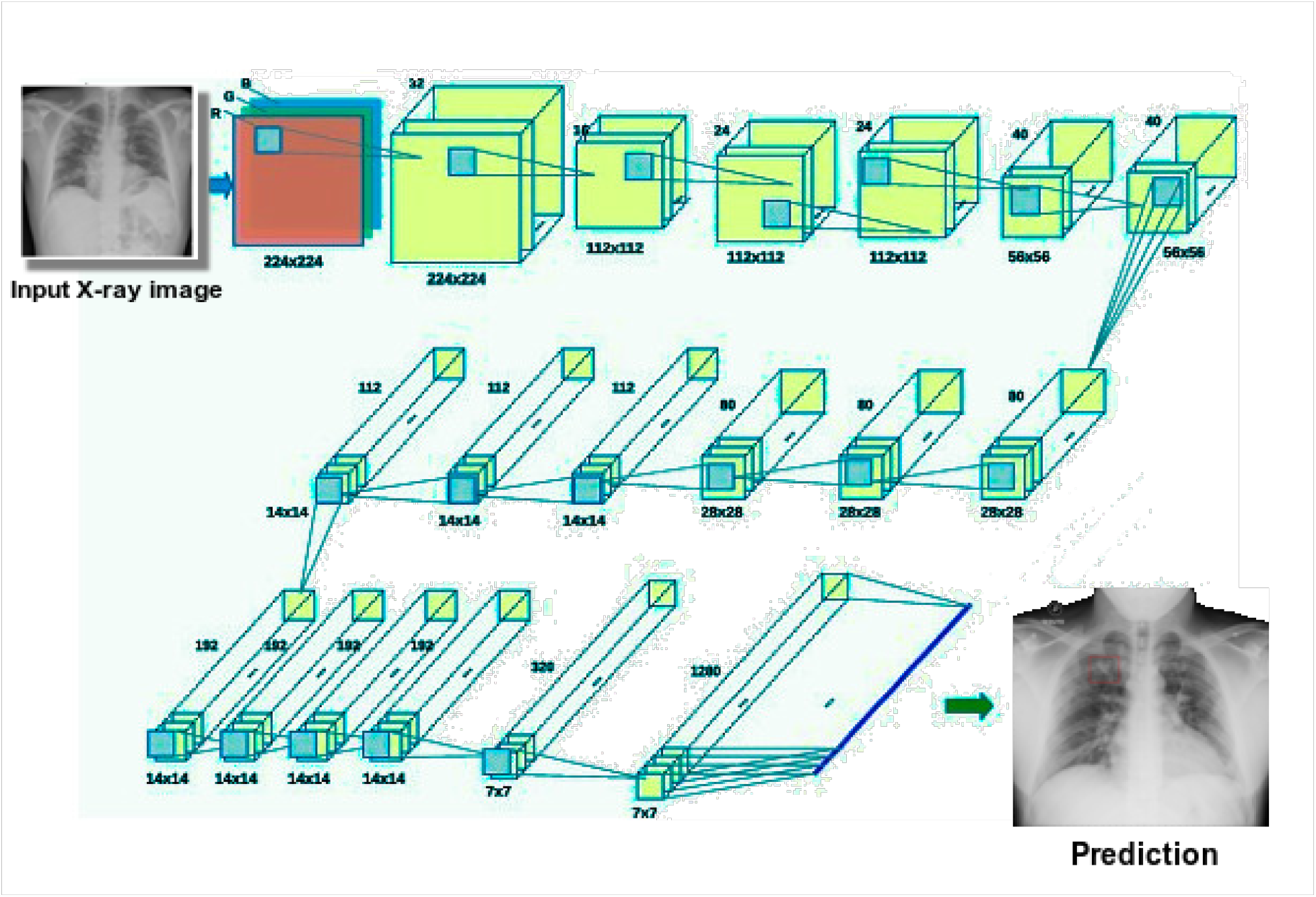
COVIDPEN: Transfer learning on pruned EfficientNet-B0 based model for COVID-19 disease identification.

In this paper, we make use of pre-trained ImageNet weights in EfficientNet-B0 model and perform transfer learning on the disease^1^ classifier. Our proposed model COVIDPEN - Transfer learning (TL) on pruned EfficientNet-B0 model shown in Fig. 1. Our proposed model builds upon the original EfficientNet [6] architecture representing a intensive configuration in Fig. 1. It consists of 18 convolutional layers where each layer is equipped with a filter of dimensions (1,1), (3,3) and (5,5). The input to the model is a chest X-ray / CT image consists of three color channels (R, G, B) of which each of them having size 224×224. Each convolutional layer having corresponding filters to output a feature map which can be computed by convolving the input feature map and the kernel. The next layers are down-scaled to minimize the size of feature map. The feature map are kept equal in size. Also, the second convolution layer is made of 16 filters and then next layer of 24 filters. However, the entire number of filters is of 1280 depth for the last layer which then fed to the fully-connected layer. Such an increasing number of convolution layer with proper depth allows to inherently capture complex features [6]. And, a kernel with larger size keeps high-resolution patters, whereas a small size kernel is better for extraction of low resolution patterns. We then adopt transfer learning on the pruned EfficientNet-B0 model, where it outputs the biases and weights and is used to classify the inputted frontal-view chest X-ray or CT images.

### C. One Cycle Policy

We prefer one cycle policy over global learning rate with decay due to earlier convergence mechanism. One cycle policy was initially introduced in [26] for stochastic gradient descent (SGD) and it differs from cyclical learning rate in a way that the extent of learning rate for its minima and maxima can be fixed with a step size. It allows the model to attain better accuracy with a faster convergence. This reward policy approach benefits both the simulated annealing and curriculum learning which are well-known used in deep learning.

We report the progress of learning rate and momentum during one cycle policy in Fig. 2 of our proposed model. In plot, the number of iterations depicts the step size, where it forms a cycle made of multiple steps, and one among with the learning rate steeply decreases and for the second step in which it steeply increases. In this policy, the total number of iterations is always higher than the cycle due to monotonically decreasing orders of magnitude less than the warm-up learning rate for the remaining iterations. When learning rate reaches the critical point (maximum bound), at the same time, momentum starts to descend from 0.954 to 0.846 linearly. Deep neural networks, especially pretrained models manifest varied levels of information in their layers, which initially emanates from initial layers to learn specific features to the final layer learning domain-wise high-level features. So, it depends on the layers of the network which needs different learning rates to be fine-tuned upon some task.

**Fig. 2:**
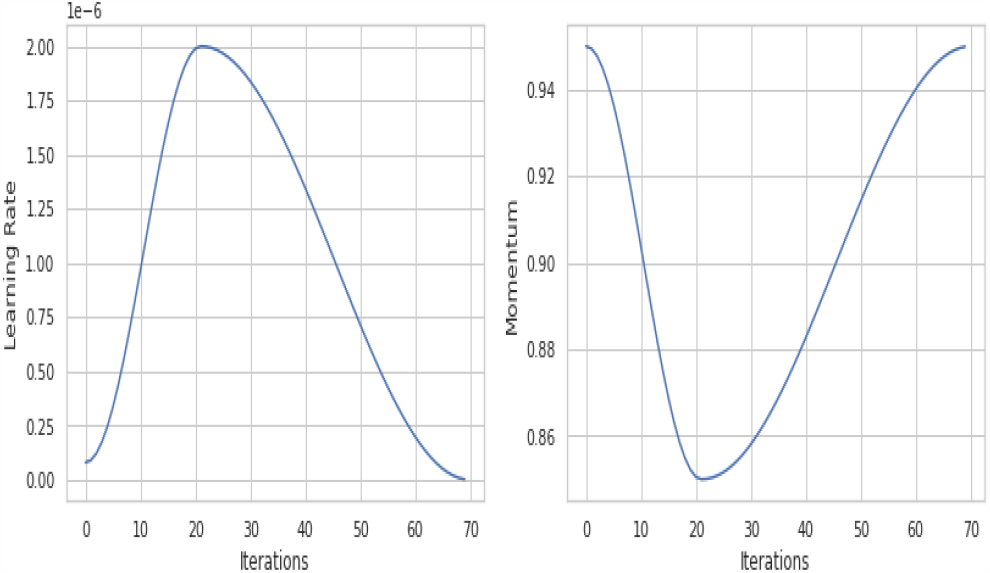
Learning rate and Momentum plot based on the One Cycle Policy.

## IV. Experiment

### A. Dataset

We use two kinds of datasets - COVID Computed Tomography (CT) scans [27] and chest radiographs collections [28] with labels delineating the clinical findings in the corresponding datasets. The chest CT scans of COVID-19 and non-COVID-19 samples were collected from several published articles in Journals. This dataset constitutes two kind of samples categorized (or labelled) as COVID-19 and non-COVID-19 with over 746 image samples. In combination, we split the dataset for training steps of our proposed models and other baselines which contains 75% of images for training and rest for test set. The resolutions of CT scans samples in terms of image size and pixels are diverse. The width*×*height range of image samples are between 124*×*153 and 1,458*×* 1853. The train and test splits comprises of both positive and negative samples of COVID-19 CT scans which is kept for conciseness. A visualisation of CT scans after augmentation is shown in Fig. 4. We demonstrate the proposed model effectiveness on the second dataset which contains a detailed diagnosis information of labelled chest radiographs image samples. This dataset contains five classes categorized as *Normal, Bacterial, Tuberculosis, Viral*, and *COVID-19* due to their generic differences in radiologic and clinical features. To diagnose COVID-19 cases, we sampled other classes apart from COVID-19 as non-COVID for the detection of coronavirus disease. Overall, the entire chest radiographs dataset were randomly splitted into train and test sets with the ratio of 0.8 and 0.2. We report a visualisation of this dataset after augmentation in Fig. 3.

**Fig. 3:**
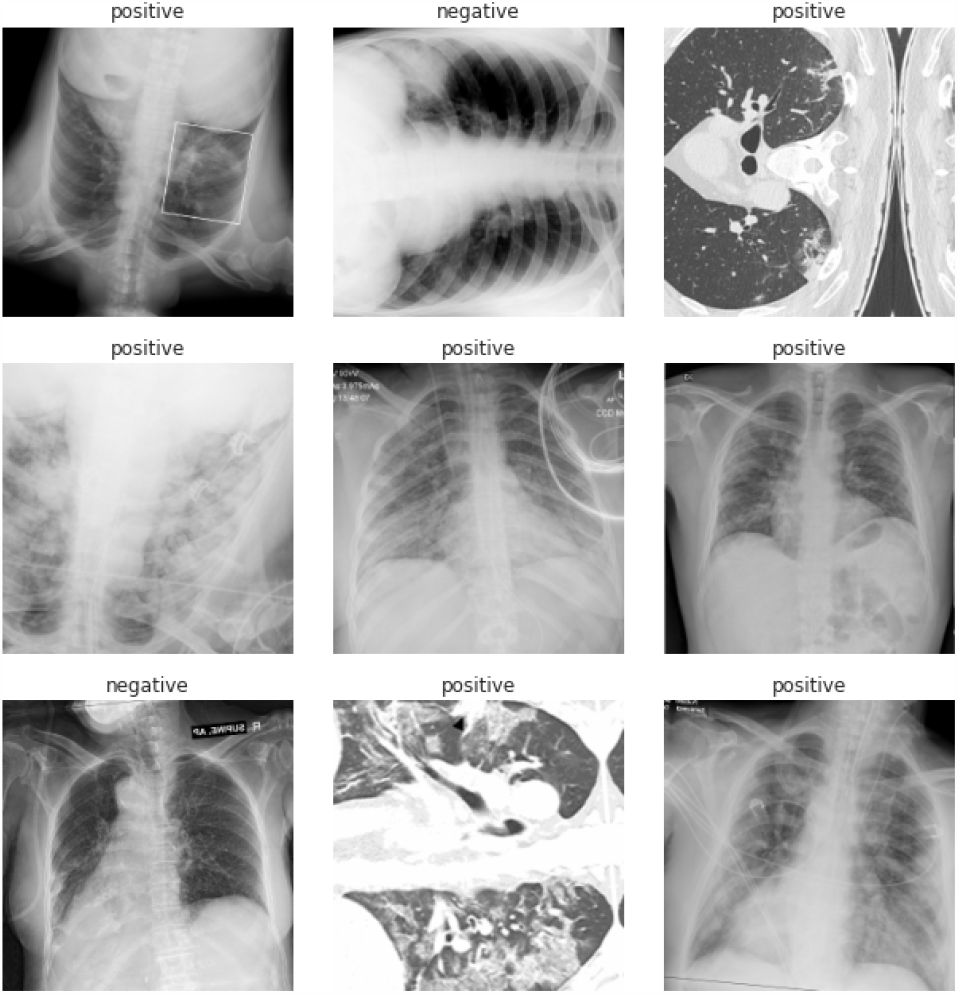
COVID-19 Chest X-ray Sample.

**Fig. 4:**
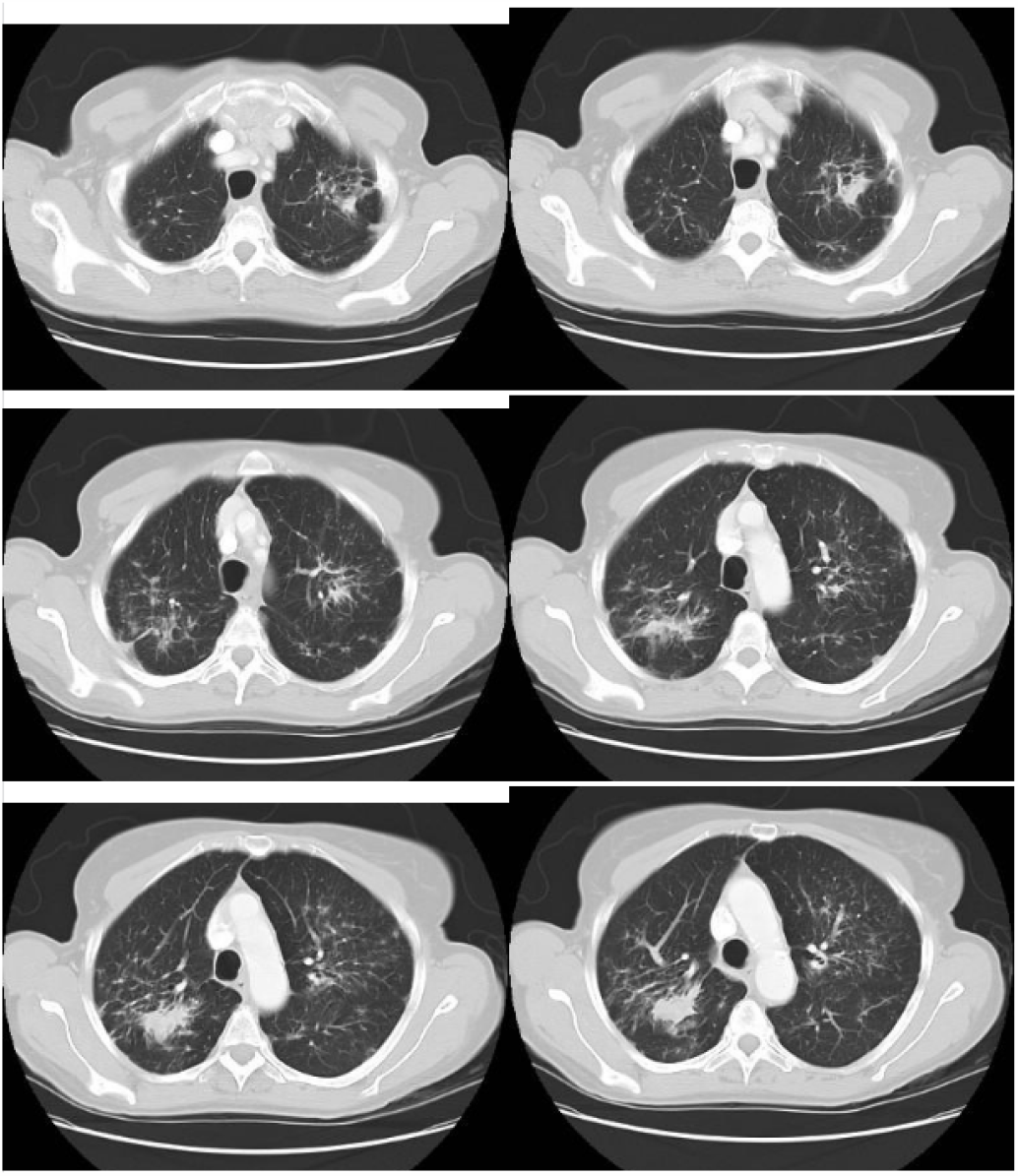
COVID-19 Computed Tomography Scans Sample.

**Fig. 5:**
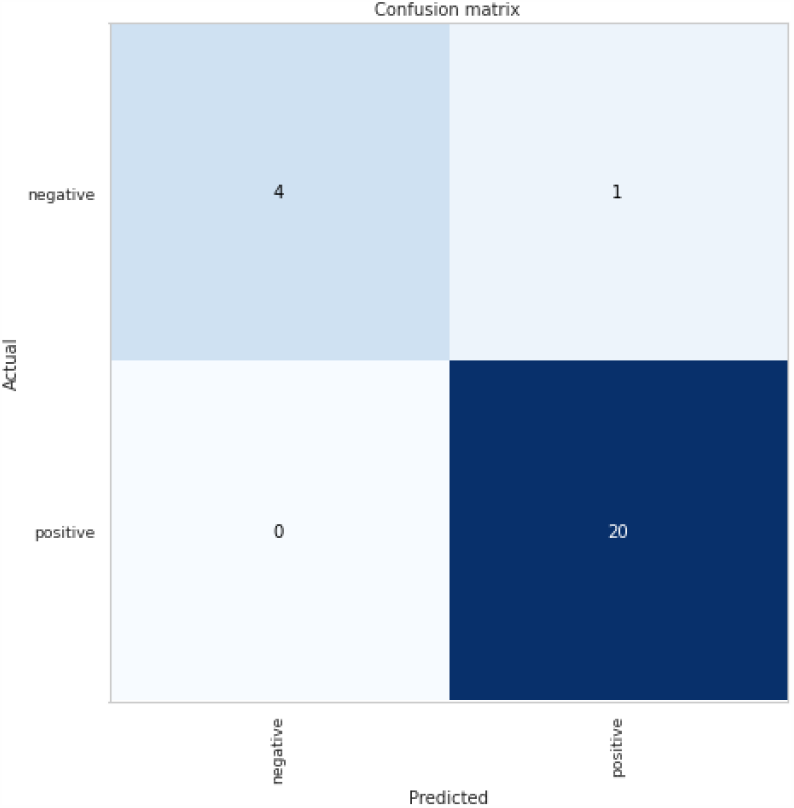
COVIDPEN Confusion Matrix.

We detail the augmentation method and steps in IV-C.

### B. Training

In this section, we report a detailed analysis of the training steps of our proposed model with parameter settings. The proposed model uses the pre-trained weight of ImageNet for the network to initialise which then trained using the chest radiographs and CT scans dataset. A detailed analysis of EfficientNet-B0 model [6] when trained with original architecture, on fine-tuning and then transfer learning mechanism. We found the transfer learning mechanism on EfficientNet-B0 model makes the training stable even on a limited labelled data. For training optimization, we use Adam optimizer at a learning rate of 1e*−*4 which is trained for over 30 epochs. The training steps make use of a learning rate scheduler (ReduceL-ROnPlateau) at alpha of 0.2 based on validation performance metrics. The batch size of 32 was used for all baselines and the proposed model. We make use of one cycle policy and a detailed overview is reported in Section III-C. However, a learning rate of 2e*−* 6 were used to train on for 5 epochs after loading the trained model weight which was trained for over 30 epochs. To minimize overfitting, we make use of L2 regularization as it is a common problem for using deep neural networks on limited data, in particular for image datasets.

### C. Test Time Augmentation

We use test time augmentation to improve the test accuracy. TTA is used for data augmentation on a test set of images to generate several variety of it and average the prediction for them. There are several image classification tasks such as employed TTA for cell segmentation of microscopy images. Several transformations such as horizontal / vertical flipping, cropping, rotation, lightning, zooming, scaling, warping, etc. can be applied to form a concrete augmentation of image dataset. In our work, we apply some of the transformations during test-time augmentation which includes flipping, vertical flipping, lightning by 5%, zooming by 10%, and 10% of warping. Also, we normalized the augmented data using ImageNet transfer learning after the final transformations. TTA based transformations shows improvement for some of the CNN-based models such as variants of EfficientNet and ResNet. However, for some of the models TTA prediction improved a bit and descends for some. We report the TTA predictions in Table II. We used test-time augmentation to ensure that it resembles cross-validation scores which is a reliable measure of how well neural networks perform on unseen image samples.

**TABLE I:** COVID-19 Datasets

| # Images | Computed Tomography | Chest X-ray |
| --- | --- | --- |
| Train | 596 | 246 |
| Test | 150 | 66 |

**TABLE II:** Evaluation Results: Comparison of baseline models where cell coloured in yellow is for the best model in corresponding evaluation measures. The Chest X-ray dataset columns are shaded in gray and CT scans dataset are shaded in cyan.

| Metric<br>Model | Chest X-ray |  |  |  |  |  | Computed Tomography (CT) |  |  |  |  |  |
| --- | --- | --- | --- | --- | --- | --- | --- | --- | --- | --- | --- | --- |
|  | Evaluation Metrics |  |  |  |  |  |  |  |  |  |  |  |
|  | Accuracy | Precision | Recall | F1 | AUROC | TTA | Accuracy | Precision | Recall | F1 | AUROC | TTA |
| VGG19 | 0.91 | 0.91 | 0.93 | 0.92 | 0.92 | 0.86 | 0.69 | 0.73 | 0.79 | 0.76 | 0.71 | 0.70 |
| ResNet34 | 0.69 | 0.72 | 0.89 | 0.80 | 0.75 | 0.76 | 0.77 | 0.81 | 0.86 | 0.83 | 0.78 | 0.78 |
| ResNet50 | 0.91 | 0.92 | 0.95 | 0.94 | 0.94 | 0.90 | 0.84 | 0.80 | 0.89 | 0.84 | 0.87 | 0.84 |
| DenseNet201 | 0.88 | 0.95 | 0.88 | 0.91 | 0.95 | 0.96 | 0.74 | 0.79 | 0.91 | 0.85 | 0.80 | 0.80 |
| EfficientNet-B0 | 0.83 | 0.87 | 0.87 | 0.87 | 0.83 | 0.89 | 0.74 | 0.80 | 0.88 | 0.84 | 0.76 | 0.79 |
| Fine-tune EfficientNet-B0 | 0.89 | 0.88 | 0.88 | 0.88 | 0.91 | 0.90 | 0.81 | 0.79 | 0.85 | 0.82 | 0.83 | 0.82 |
| **COVIDPEN (Ours)** | **0.96** | 0.92 | 0.96 | 0.94 | 0.92 | 0.93 | **0.85** | **0.81** | 0.92 | 0.86 | 0.84 | **0.85** |

### D. Performance Measure

To evaluate our proposed model and comparable baselines, we use several metrics such as test accuracy, precision, recall, F1 measure, and area under the probability curve - ROC. A detailed overview of the evaluation metrics is as follows:

- *Accuracy*: The accuracy here refers to the test accuracy i.e., accuracy on hidden / test samples.
- *Precision*: Precision refers to the positive predictive value and is the ratio of true positive samples to the samples containing true and false positives.
- *Recall*: This measure is the most well-known metric to test the effectiveness of a classifier. Recall, also refers to Sensitivity or true positive rate which represents a classification model discards a positive prediction.
- *F1*: Similarly, a well-known measure for several kind of tasks related to machine learning problems, in particular, classification. It is the harmonic mean between precision and recall estimations.
- *Area under the receiver operating characteristic (AU-ROC)*: The AUC metric is one of the most common measure which sum-up the information contained within the ROC curve, which depicts sensitivity versus false positive rate at some certain thresholds. A better AUC score falls under the higher values to differentiate between COVID and non-COVID images. We report AUROC score in Table II for each and every model which includes baselines as well.

All of the measures are used to report an extensive evaluations across different baseline models in Table II.

### E. Results

In this section, we report the experimental results i.e., the COVIDPEN performance and an extensive comparison with CNN-based baselines across varied evaluation measures. A detailed result is reported in Table II The main metric measures are test accuracy and test-time augmentation scores as it correctly classifies the cases into each label - *positive* for COVID-19 and negative for *non-COVID-19*. We employ seven CNN-based models which are VGG19 [23], ResNet34 [24], ResNet50 [24], DenseNet201 [25], EfficientNet-B0 [6], finetuned EfficientNet-B0, and COVIDPEN. For chest X-rays dataset, we found that based on the accuracy, apart from the proposed model, VGG19 and ResNet50 performances are equal and shown to be second best model. However, we performed test-time augmentation to see which one stands best among themselves, and then TTA prediction for ResNet50 is 90% which shown to be effective than VGG19, but there is almost no-difference betweeen ResNet50’s accuracy and TTA score which is the reason why it was better than VGG19. In other case, based on the TTA score, DenseNet201 performance is shown to effective among all models.

For CT scans dataset, we found ResNet50 to be effective after our proposed model. However, for this dataset, our proposed model performances based on test accuracy and TTA score shown to be among the highest.

## V. Interpretability

In this section, we investigate the apparent benefits of our proposed model with explainable approach to describe the prediction outcome. As this allows healthcare organisations to benefit patients by ensuring responsible predictions made by our proposed model. The idea of explainability stems from [29] to provide accountability and transparency of classification models. The deep neural network is a black-box which makes difficult to explain its predictions. In terms of medical imaging datasets, especially COVID-19 chest X-ray and CT scans to which interpretability is of utmost importance. We can see from the saliency maps shown in Fig. 6 and Fig. 7 of region (or patch) importance that the bacterial features (coronavirus or other non-COVID diseases) have the greatest influence on the result of the prediction in the selected model is the mean area, followed by mean concave region and mean texture.

**Fig. 6:**
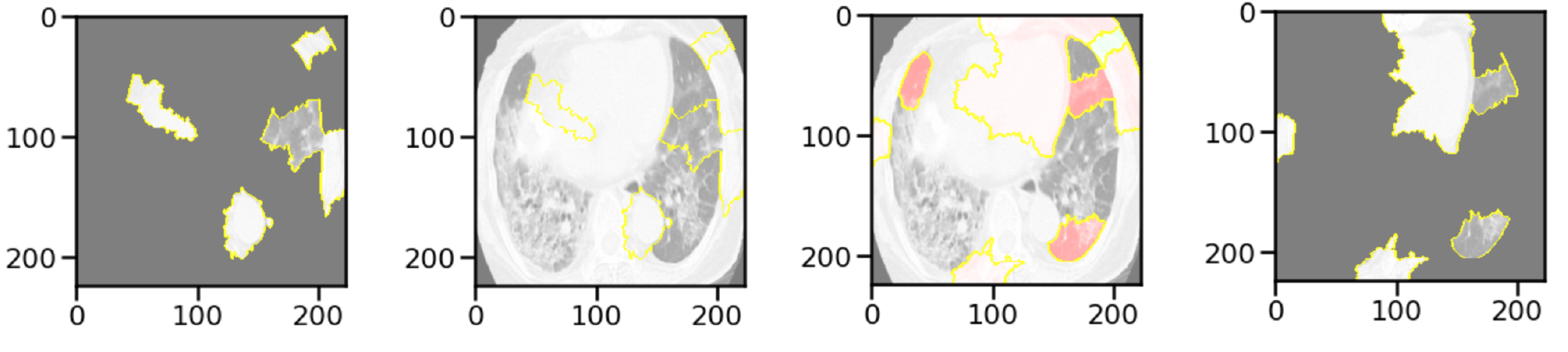
Model Interpretability - COVID-19 CT Scan Dataset.

**Fig. 7:**
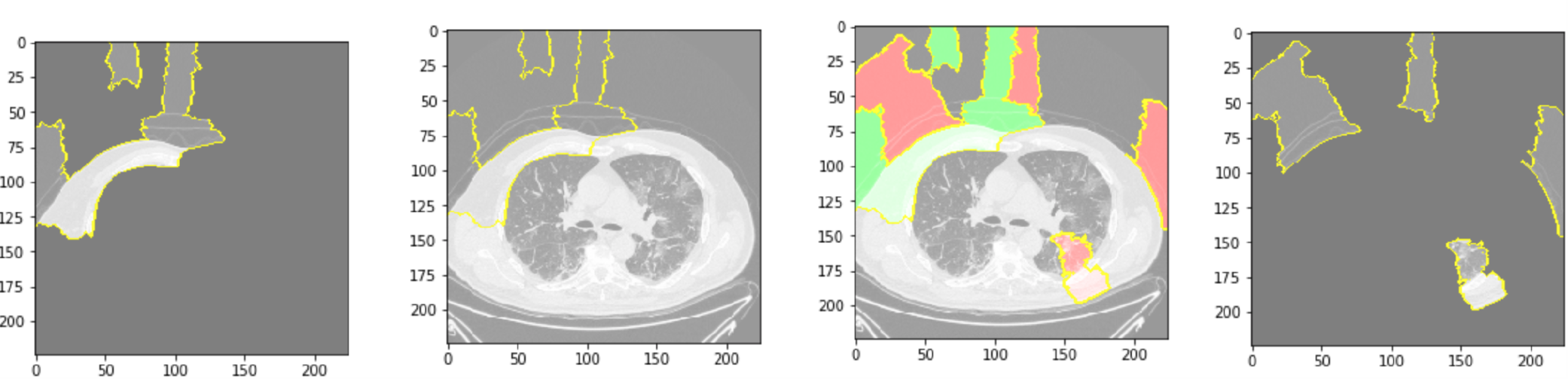
Model Interpretability - COVID-19 Chest Radiographs Dataset.

We employ local interpretable model-agnostic explanations (LIME) [29] for the interpretability of the predictions made by our proposed model. The trained model weight were the input to LIME explainer which perform perturbations on the features (at pixel level) over a set of predicted samples (we have taken 200 samples which is shown as plotted over the axes in Fig. 6 and Fig. 7) on test set of both datasets, and it interprets the weight resulted after training our proposed model - COVIDPEN at the local region in the feature space. The masks around images are generated by segmenting the predicted sample (CT scan in Fig. 6 and Chest X-ray in Fig. 7) into superpixels and superpixels then inspected based on the predictions values. Superpixels are interconnected pixels composed of similar colors and pixels can be lightened based on the user-defined color such as gray. However, in our case the mask boundaries are in yellow color. The exposed interpretability is an informed feature engineering driven by some inconsequential features which can be understandable to medical experts. We segment such behavioural features by removing and in-painting textual regions. The interpretability plots are the top two images from our prediction set made by the TL EfficientNet-B0 classifier. The regions shaded in pink and green detect superpixels that contributed to and against prediction of chest radiographs and CT scans.

The model explainability is a part of post-hoc analysis on the trained model to ensure predictions feasibility in real scenario for healthcare providers.

## VI. Conclusion and Future Work

In this work, a deep neural network-based classifier is proposed i.e., COVIDPEN to diagnose COVID-19 and non-COVID-19 cases from chest radiographs and chest X-ray datasets. We begin with the initial baseline models composed of neural network based architectures which can be adopted to detect or classify COVID-19 cases. Our proposed model based on EfficientNet is shown to perform better based on the prediction using accuracy and test-time augmentation measures. The granularity of predictions from our proposed model is on par among other DNN based models. We further show that our predictions are interpretable using LIME framework. It also describe the implications of our proposed model thoroughly.

In future, we intend to scale experimentation on a larger dataset for expliciting the capability of our proposed model. Based on our results, it signifies that architectures such as EfficientNet and EfficientDet^2^ could be leveraged to assist radiologists in the CT or chest X-ray mediated diagnosis of COVID-19 cases.

## Data Availability

This work uses publicly available datasets.

## Footnotes

1 Disease in this paper refers to COVID-19 symptoms, however we do not use any external data (other bacterial disease such as Pneumonia) to train over

2 https://github.com/google/automl/tree/master/efficientdet

